# Stalled malaria control – root causes and possible remedies

**DOI:** 10.1101/2022.07.09.22277454

**Authors:** J. W. Hargrove, G. A. Vale

## Abstract

**Background:** Malaria control has been stalled for some years in many African countries. We suggest reasons for the stalling, and ways of remedying the situation.

**Methods:** We analyse malaria data from Kenya and Tanzania using mathematical analysis and a deterministic model for mosquito and malaria population dynamics. The model was produced in Microsoft Excel and is usable by persons who are neither mathematicians nor specialised modellers.

**Results:** In Kenya, there was no significant decline in malaria incidence during the last decade, despite 50-80% of the human population owning and using insecticide-treated bed-nets (ITN). Similar situations exist in Tanzania and Uganda. There were only limited declines in malaria incidence in Kenyan counties where indoor spraying of residual insecticides covered about 90% of the dwellings. This is especially surprising since, in the earlier decade, a rapid drop in malaria incidence followed much less intense control operations. While there have been well documented increases in resistance of anopheline mosquitoes to various pyrethroids, these are not sufficient to explain the stalled control. Instead, we suggest that this is largely due to a change in the species composition and behaviour of the vector populations, consequent on the widespread use of ITN. Quantitative support for this view is offered by our mathematical analyses and modelling of published data on changes in malaria incidence and levels of access to, and use of, ITN. The modelling also suggests that a resumed decline in the incidence of malaria might best be achieved by increasing ITN coverage as close as possible to 100% and supplementing this effort with limited application of control measures that kill mosquitoes attempting to feed off non-humans. Particular attention deserves to be given to the insecticide treatment of cattle hosts and the refinement of artificial baits for outdoor deployment.

**Conclusions:** Current levels of indoor residual spraying (IRS) and ITN will not result in any significant improvement. If, however, ITN ownership and effective use can be increased closer to 100%, modest levels of additional control outdoors should result in substantial reductions in incidence. Treatment of adult mosquitoes visiting livestock is a promising option.

## Background

Following the roll-out of ITN between 2000 and 2010, there were dramatic declines in malaria incidence in much of Africa (Bhatt et al., 2015). Since then, however, malaria control has stalled in several African countries, including Kenya, Tanzania and Uganda (World Health Organisation, WHO, 2021a). Many different explanations have been offered as to why the control has stalled. For example, it is claimed that there has been inadequate commitment on the political, financial and social fronts (WHO, 2021b). Other explanations single out the development of insecticide resistance by anopheline mosquitoes (Hemingway et al., 2016; Churcher et al., 2016; Ranson & Lissenden, 2016; Kleinschmidt et al., 2018) or, more generally, reductions in the quality and longevity of the bed nets (Ochomo et al., 2013; Wills et al., 2013; Lindsay et al., 2021).

It has also been suggested that heterogeneities, and changes, in human behaviour limit the effective exposure of mosquitoes to the nets, particularly in situations where the use of indoor residual spraying (IRS) can encourage the belief that net use is of reduced importance (Protopopoff et al. 2018). Finally, there are indications of an altered abundance of the strains and/or species of mosquito responsible for transmitting malaria (McCann et al., 2014; Mustapha et al., 2021). This can lead to changes in the times and places at which the mosquitoes feed, and hence reductions in the chances of the insects being killed by the nets (Mawejje et al., 2021; Sangbakembi-Ngounou et al., 2022).

Each of the explanations for the stalled control of malaria is likely to have been applicable, at least to some extent in some countries for part of the time. Moreover, several of the explanations have led to actions presumed capable of overcoming the stalling. For example, in response to the development of resistance to the pyrethroids, new long-lasting insecticidal nets (LLIN) – impregnated with more than one insecticide or an added synergist – have been shown to be more effective than those dosed only with a pyrethroid (Tiono et al., 2018; Lynd et al., 2019; Staedke et al., 2021; Mosha et al., 2022), as have such nets used in conjunction with IRS (Katureebe et al., 2016; Protopopoff et al., 2018). In general, these actions have been associated with welcome reductions in the incidence of malaria, but they have fallen far short of eradicating it.

Efforts to kill anopheline mosquitoes outside of houses have not been pursued with as much vigour as the efforts to improve the performance of ITN and IRS. A study in Ethiopia did, however, demonstrate that *An. arabiensis* fed readily on cattle treated with deltamethrin and were killed when they contacted recently treated animals (Habtewold et al., 2004). This has been followed by suggestions, backed by modelling studies (Iwashita et al., 2014; Franco et al., 2014; Yakob et al., 2017), that insecticide treated livestock (ITL) could be useful in malaria control. The approach has already been attempted in Pakistan (Rowland et al., 2001) but there has been no large-scale trial in Africa.

The use of attractive toxic sugar baits (ATSB) has, likewise, been suggested as a complement to the use of bed nets (Gu et al., 2011; Beier et al., 2012). In an experiment in Mali, Traore et al. (2020) demonstrated large reductions in mosquito population density, the numbers of older, and sporozoite infected, females and in the entomological inoculation rate (EIR). While this is a promising development, there has been no convincing demonstration in Africa of the use of ATSBs in achieving sustained reductions in malaria incidence.

Historically, malaria has often been successfully controlled in Europe and north America through larval source management (LSM) – killing the immature stages, either by using chemicals or by physically removing the aqueous environment required for larval development (see Tusting et al. (2013) for a review of this approach and references). There have been suggestions for ways in which LSM could be carried out in Africa (Afrane et al., 2016; Ingabire et al., 2017) but as yet there has been no demonstration of a sustained effect on malaria incidence.

Williams et al. (2018) provide a more comprehensive list of vector control methods that could be used to complement the impact of LLINs and IRS on malaria incidence, but conclude that, at the time of their publication, the quality of the evidence is greatest for LSM and topical repellents, relative to the other vector control tools evaluated, although existing evidence indicates that topical repellents are unlikely to provide effective population-level protection against malaria.

Pilot trials of the RTS,S malaria vaccine hold out renewed promise that a vaccine will strengthen efforts to control malaria in Africa and save tens of thousands of young lives (Adepoju. 2019). Whereas RTS,S appears to be much more promising than earlier vaccines, tests indicate that it is markedly less than 100% effective (Agnandji et al., 2011; Samuels et al., 2022;) and the view of WHO is that “New and complementary tools are needed to further drive down malaria cases and deaths, with a view to ultimately achieving the vision of a world free of malaria” (Adepoju. 2019). Moreover, even if vaccines were 100% effective and easy to use, we should still be well prepared to tackle the disease with all available techniques because the long-term efficacy of any one method cannot be guaranteed. In particular, we need to continue refining vector control as a complement to the vaccines.

Despite the indications above of the large amount of work carried out in malaria control, and the wide variety of suggested control measures, there seems to be no comprehensive and generally agreed analysis of why malaria control has stalled in many parts of Africa. Nor is there agreement on what types of new control measures, or modifications of existing methods, might best overcome the stalling. The present work involves the combination of a literature review, and modelling of the dynamics of mosquito populations and malaria transmission, leading to a suggested design for a clustered randomised control trial (CRCT) of what we identify as the most promising means of overcoming the stalled control.

## Methods

### Literature search and mathematical analysis

We collated information from the literature to show the general pattern in which malaria control became stalled during the use of ITN in Tanzania, and ITN and IRS in Kenya, from 2000 to 2021. We then explored simple mathematical approaches to see how well the time series of malaria incidence in this period might be explained by changes in the pattern of malaria transmission, and to assess the timing and extent of any such changes.

### Model of mosquito control and malaria incidence

Given that the above analysis was consistent with there being a switch between two patterns of transmission cum control dynamics, we needed a model to help judge the extent to which the two patterns could be characterized by differences in mosquito behaviour and/or insecticide resistance. The deterministic model we created for this, termed the “M” model, was produced in Microsoft Excel, with attention to making it usable by persons who are neither mathematicians nor specialised modellers. It is available in medRxiv (Reference) and is merely outlined here.

#### Mosquitoes

Only female mosquitoes were considered. They transferred to different compartments from day to day. These were grouped into: (i) aquatic compartments, covering the combination of eggs, larvae and pupae at various days from deposition as eggs, and (ii) adult compartments which were distinguished according to days from emergence, up to a maximum of 100 days, and by the stage of their gonotrophic cycle. This cycle normally lasted for three days if there were no restriction to feeding success (Lines *et al*., 1991). Daily survival rates, fecundity and feeding success were allowed to vary with age, being low but rising steadily for young adults <10 days old, and then remaining level until decreasing steadily when adults became >50 days old. The standard carrying capacity was set at 25,000 adults per km^2^, with about 30 times as many aquatics. The high relative abundance of aquatics is consistent with the allowance that the fecundity of mature females was 40 female eggs per gonotrophic cycle at carrying capacity (Okal et al., 2015), and that the peak emergence of surviving aquatics occurred from 15-20 days after deposition as eggs (Bayoh & Lindsay, 2003). Daily death rates of aquatics and adults declined linearly with declining population density, and rates of fecundity and feeding success of adults increased linearly with declining density. The inputs for these processes meant that if the density of all population components were instantly reduced to 0.1% of carrying capacity, followed immediately by population growth with no seasonal setbacks, then the population density would return to ∼84% of carrying capacity in one year. Despite any density dependence of death rates, virtually no adults lived more than a month from emergence.

#### Hosts and infection

Two types of hosts were considered: humans that were potentially susceptible to infection by the malaria parasite, and non-humans that were never susceptible. Humans were assumed to occur at 420 per km^2^, roughly simulating their density in W Kenya (Rutto et al., 2018). The probability of a susceptible human becoming infected, after being bitten by a mosquito with a mature infection, was 95% and the incubation period in humans was 12 days (Bayoh & Lindsay, 2003). Infective humans were allowed to remain so for 20 days, after which the daily probability of getting a curative treatment rose steadily to a maximum of 10% per day at 40 days. After beingcured the humans remained uninfectable for a further 20 days, transferring thereafter to the group of re-infectable humans, at a rate rising steadily to 10% per day at 30 days. The probability of a mosquito being infected after feeding on a human with a mature infection was 95%, and the incubation period in the mosquito was 11 days (Bayoh & Lindsay, 2003). Once infected the mosquito was taken to stay so for the rest of its life. At the start of each flight season the percentage of infective mosquitoes was virtually zero, since most of the population consisted of recently emerged adults with insufficient time to acquire and mature an infection. In the middle of the flight season the percentage of infective mosquitoes was never more than a few percent since most flies did not live long enough to mature any infection they received (Lines *et al*., 1991). Virtually no young flies were emerging at the end of the flight season, so that the adults present then were mainly very old. These mosquitoes had infections rates >10% because they had had many opportunities to become infected. However, the epidemiological impact of these flies was slight because they were so few.

#### Two main scenarios

We recognised two sets of background conditions to suit different sorts of mosquito with distinctive behaviour, and hence distinctive availabilities to some of the control methods. Set 1 was intended to cover species such as *An. gambiae* ss and/or *An. funestus* whose behaviour is commonly anthrophilic and endophagic (Orsborne *et al*., 2018), and which feed mainly late at night (Cooke *et al*., 2015) when most humans are generally in bed. For these species it was taken that humans contribute 75% of their diet, and that 75% of the humans were making effective use of any nets they owned. The notion of effective use is the product of the probabilities that: (i) any owned net is properly deployed above the owner’s bed, as against, say, being stored in a cupboard, and (ii) the owner is indoors and in bed at the time when mosquitoes are trying to feed, as against say, walking back from a night visit or sitting in a chair at home. Allowing that roughly 87% of owned nets are properly deployed (Bertozzi-Villa *et al*., 2021), the 75% use of nets adopted for Set 1 implies that around 86% of the owners are indoors and in bed at feeding time. Set 2 was intended to cover species such as *An. arabiensis*, which are largely zoophilic and exophagic (Orsborne *et al*., 2018), and feed at relatively wide times of day (Cooke *et al*., 2015). For such species it was taken that humans formed only 25% of diet, and that only 25% of the owned nets were in effective use.

#### Seasonality

Adult rates of birth and death, and the rates at which the aquatics entered a dormant stage, or emerged from it, were varied seasonally in accord with the long and short rainy seasons in W Kenya. Hence the simulations involved one long and one short flight season per year, lasting ∼200 days and ∼100 days, and starting in March and November, respectively. The inputs for seasonality were the same from year to year. They were also the same for *An. gambiae* and *An. arabiensis* so that in the absence of control the simulated seasonal abundances of the two species were similar (Fig. 1). However, since *An. gambiae* was more anthropophilic than *An. arabiensis*, the former species was associated with the greater incidence of malaria (Fig. 1). For both species, the simulated incidence was low after the end of the flight seasons, especially the long flight season, and rose soon after the new flight seasons began. This is consistent with the general pattern of clinical records in W Kenya from the Kenya Health Information System (KHIS).

**Figure 1.**
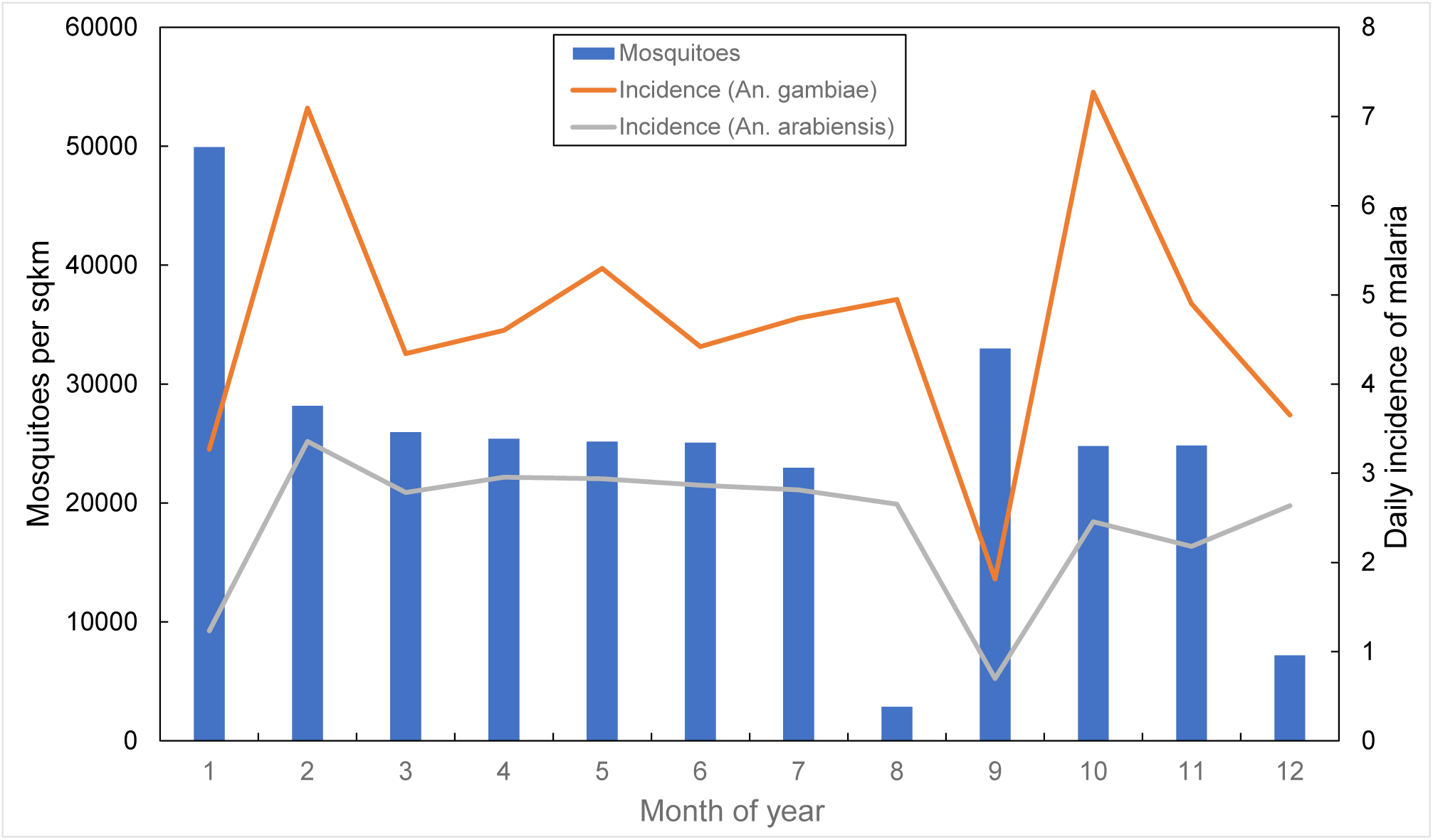
Outputs for the average daily numbers of adult female mosquitoes per km^2^ and average daily incidence of mature cases of malaria per km^2^ in each month of a year before any control. Months are counted from the beginning of the first and longest of the two flight seasons per year, so that Month 1 is March. The data for the number of mosquitoes per km^2^ apply to either *An. gambiae* or *An. arabiensis*. However, the two species produce markedly distinctive incidences of malaria because they differ greatly in the degree to which they feed on humans, as indicated in the text.

#### Control methods

While the model allows for a variety of control methods, the only ones considered in present work were: (i) bed-nets, with or without insecticide treatment; (ii) indoor residual spraying, which kills mosquitoes that rest on sprayed surfaces after taking a meal; iii. measures applied at breeding sites, to kill immature mosquitoes before they emerge (Shelley & Aziz, 1949), and (iv) killing adults feeding on non-humans, as for example by spraying insecticide on cattle (Njoroge *et al*., 2017). Unless stated otherwise, all control was imposed on a previously stable and uncontrolled population, and continued every day for five years, with steady parameter values for the efficacy of control. The impact of control was then measured in the fifth year, as the average daily values for the abundance of adults and the number of newly matured infections, expressed as percentages of the daily averages in the year prior to the start of control.

#### Simplification

We acknowledge that the M model is, like any other, a simplification of reality and, in common with other malaria models, is short of reliable values for many of the input parameters ideally required. For example, although it is widely recognised that density dependent processes affect the population dynamics of malaria vectors (Muriu *et al*., 2013), we could find no satisfactory field information for how population growth is governed by the combination of density dependencies in the rates of birth, death and feeding success. There are not even tight indications for the absolute densities of the vector populations (Costantini *et al*., 1996). Moreover, the available indications for the feeding behaviour of mosquitoes vary substantially within and between different mosquito species (Orsborne *et al*., 2018), and they bear an uncertain relationship to human activity (Sherrard-Smith *et al*., 2019). Such problems become especially severe in places such as E Africa where as many as 17 species of mosquito can be sympatric, with limited information for how the dynamics of the separate species interlock (Ondiba et al., 2019). Given these difficulties, we cannot claim that the model produces precise simulations. However, we do suggest that simple modelling of the control of one species at a time, with focus on the effect of changing just one or a few parameters, is useful in informing a common-sense understanding of pertinent principles.

## Results

### Collation of published data

#### ITN coverage and malaria incidence in E Africa

The use of ITN in Kenya increased rapidly from around 5% in 2000-2004 to about 50% in 2009-2011 (Fig. 2A). Mean usage continued to increase after 2011, albeit with cyclical increases and decreases reflecting mass distributions of nets in 2011/12, 2014/15 and 2017/18, interspersed with deterioration in net access and use (Bertozzi-Villa et al., 2021). These authors define access as the proportion of the population that could sleep under a net assuming two people per net (Kilian et al., 2013), and define use as the proportion of the population that does sleep under a net. The use rate can then be defined as use among those with access. Malaria incidence in Kenya declined by 70% between 2004 and 2009. Most of this decline has been attributed to the use of ITN (Bhatt et al., 2015), and specific studies find that use of such nets leads to reduced infection rates and the protection of life (Pryce et al., 2018; Adetokunboh, 2021). The modelling of Killeen & Smith (2008) had predicted this effect of ITN on malaria incidence. Despite the continued increase in mean net coverage, however, national malaria incidence has not changed significantly since 2009 (Fig. 2B). Nor have the oscillations in net coverage been associated with any detectible changes in malaria incidence.

**Figure 2.**
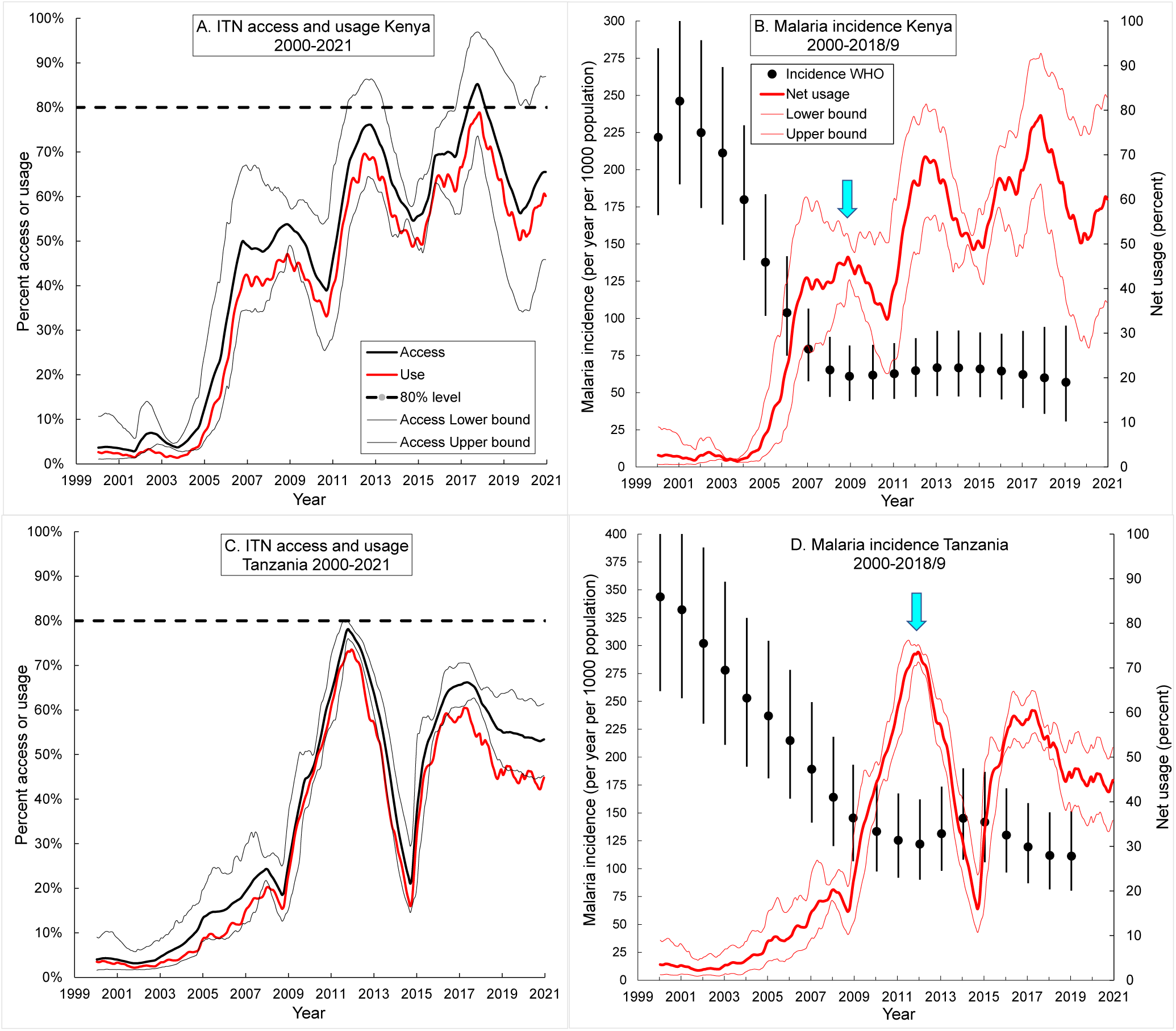
ITN access and usage from 2000 to 2020 in A. Kenya, C. Tanzania. Dotted line indicates the 80% level regarded as universal coverage (Bertozzi-Villa et al 2021). Malaria incidence from 2000 to 2019 in B. Kenya and D. Tanzania (WHO, 2021a; World malaria report, 2021), and usage of ITNs during this period. The blue arrows denote the years 2009 and 2012 when net usage hit its first peaks in Kenya and Tanzania, respectively, and malaria incidence began to plateau. Thin lines, black in A and C, and red in B and D, indicate the confidence intervals for percent access and usage, respectively.

In Tanzania, similarly, incidence declined by 65% between 2000 and 2012, during which time it was estimated that up to 73% of the population were using ITN (Fig. 2C, D). There was a significant decrease in net usage between 2012 and 2015 and a simultaneous increase in incidence – but the rapid increase in net usage thereafter made little impact on incidence. For both countries the most interesting and important result is that marked oscillations in net cover, between 2008 and 2019, were not associated with any statistically significant change in malaria incidence. The implication is that the ITN had become of reduced consequence. This is consistent with the ideas that most of the malaria transmission during that period was occurring outdoors, and/or that the mosquitoes had become resistant to the insecticide on the nets.

#### Malaria control using IRS in Western Kenya

Further evidence consistent with the above ideas is provided by the results of a concerted campaign of IRS carried out in Migori and Homa Bay counties, W Kenya (Fig. 3), between 2017 and 2020 by the United States President’s Malaria Initiative (PMI) (Johns & Haile, 2021; PMI, 2021). The adjacent counties of Kisumu and Siaya were not sprayed. IRS was carried out in Migori County in 2017 and there was a sharp decline in malaria incidence, relative to 2016 (Fig. 3). There were, however, similar declines in the adjacent counties of Homa Bay, Siaya and Kisumu, none of which was subjected to IRS in 2017. That is to say, the decline in incidence in 2017, in Migori, may have been unrelated to the use of IRS in that county. This conclusion is consistent with the data for 2018-2020 – where three years of intensive application of IRS in Migori County saw no further decline in malaria incidence (Fig. 4).

**Figure 3.**
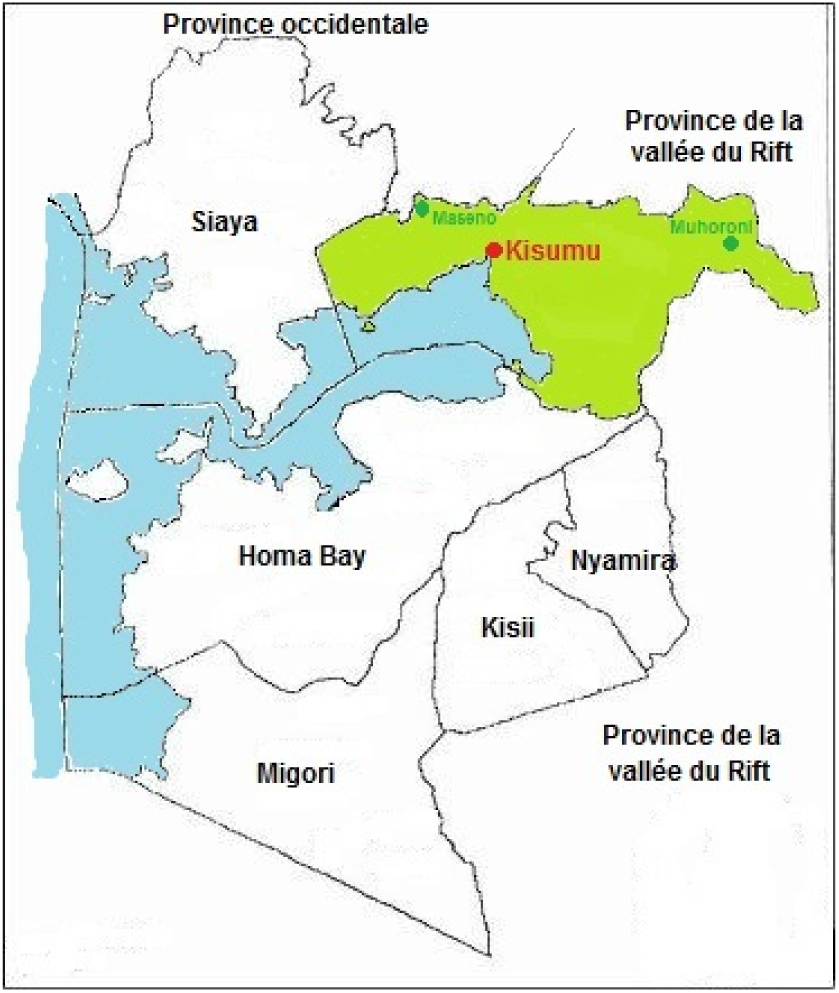
Map of Nyanza Province, Kenya, by county.

**Figure 4.**
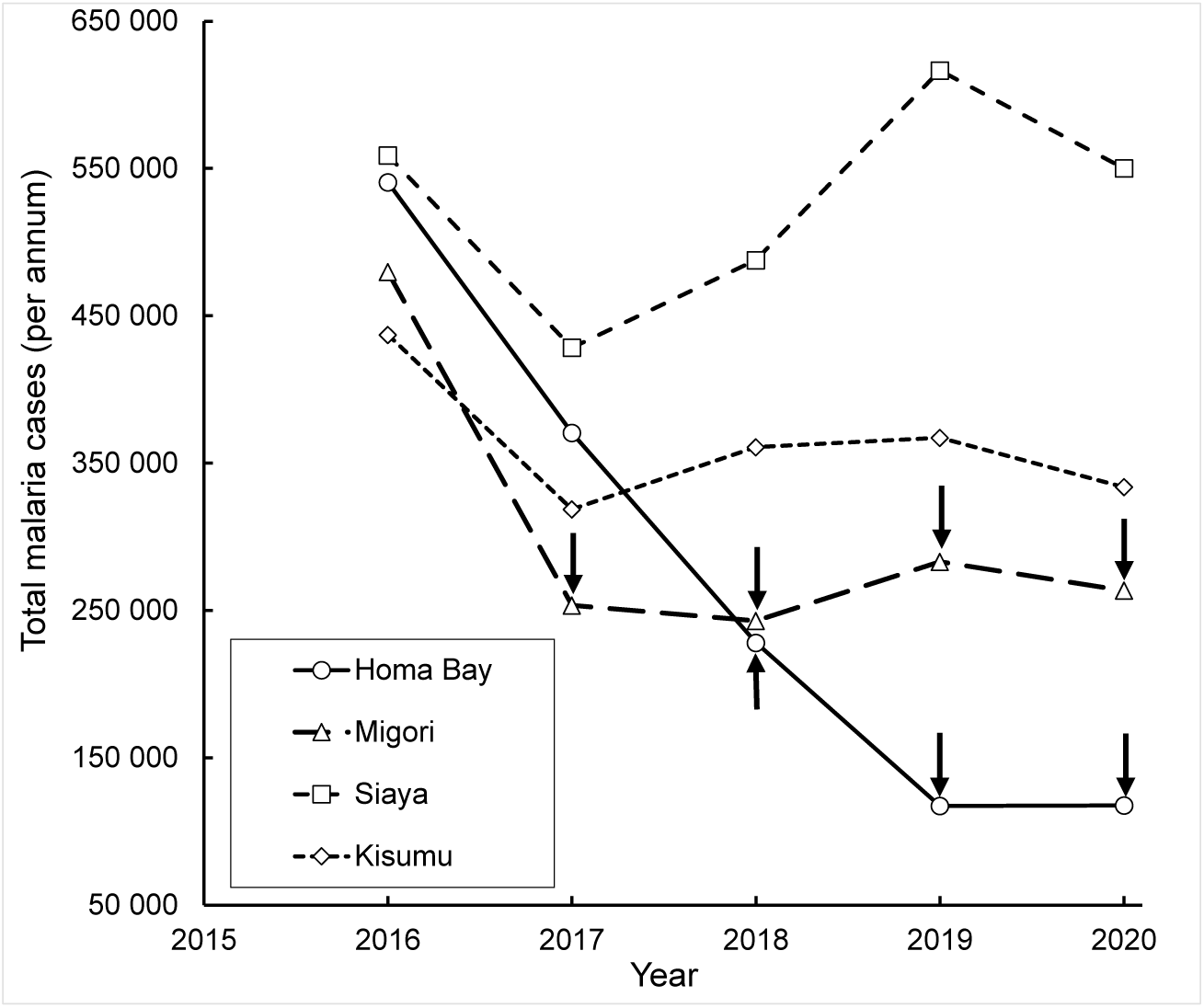
Malaria incidence *per annum* in four counties, Nyanza Province, Kenya. Black arrows indicate years in which IRS was carried out in the counties indicated.

In Homa Bay County, IRS applied in 2019 was associated with a decline in incidence which was not seen in the other three counties (Fig. 4). There was, however, no further discernible decline in malaria incidence between 2019 and 2020 in either Homa Bay or Migori counties, which recorded about 780,000 malaria cases over this period. This despite the fact that in those two years the PMI – via its *VectorLink* Project – sprayed an impressive 92% of structures in both counties, involving the expenditure of US$20 million (Johns & Haile, 2021). There was an additional unknown expenditure by the Kenya Government on the distribution of nearly 3 million insecticide-treated nets in 2017/18 (Table 1) (Government of Kenya, 2017). The declines in malaria incidence in Migori County in 2017-2020 were markedly smaller than in Homa Bay – even though Migori was sprayed in a greater number of years (Fig. 4).

**Table 1.** Numbers of insecticide treated nets distributed by county in Nyanza Province, western Kenya, 2011-2018 (Government of Kenya, 2017)

| County | Years |  |  |
| --- | --- | --- | --- |
|  | 2011/2012 | 2014/2015 | 2017/2018 |
| Homa Bay | 466 172 | 662 582 | 768 294 |
| Kisumu | 424 060 | 675 163 | 694 600 |
| Migori | 469 788 | 638 840 | 691 950 |
| Siaya | 371 866 | 641 900 | 641 900 |
| GT | 1 731 886 | 2 618 485 | 2 796 744 |

Summarizing all of the above indications for the various control campaigns in E Africa, a high incidence of malaria was sustained despite significant distribution of ITN, and the additional use – at great cost – of intensive IRS in W Kenya. These results are consistent with the suggestion that, since 2008, most of the malaria transmission occurred outdoors, and at times when humans were not in bed and so not protected by ITN. Unless it can be shown that, after 2008, malaria vectors in W Kenya were highly resistant to pyrethroids used on ITN and, simultaneously, to the organo-phosphate used in IRS, the results suggest that most of the malaria transmission occurred outdoors, and at times when humans were not in bed and so not protected by ITN. We present below further evidence pertaining to these possible explanations for the data.

#### Changes in proportions of mosquito species

Published results highlight dramatic changes in the relative abundance of mosquito species in E Africa since about 2000. For example, from 1970-1998, *An. gambiae* contributed 85% of catches of adult female mosquitoes in W Kenya – but, by 2009, its proportion decreased precipitately to 1%, at which point 99% of mosquitoes sampled were *An. arabiensis*, and samples of immature stages provide a similar picture (Bayoh et al., 2010). *An. arabiensis* is now the most important malaria vector in W Kenya (Abong’o et al., 2018) and although many of this species are found feeding off cattle, they are infected with malaria parasites at rates comparable to those feeding off humans (Abong’o et al., 2018). Moreover, Njoroge et al. (2017) found that, whereas few mosquitoes can be caught indoors in W Kenya, many can be caught outdoors using cattle-baited traps.

The indications are that the control operations were themselves the cause of this change in the species balance of mosquitoes. Thus, ITN and IRS would be particularly effective for reducing the abundance of the endophagic *An. gambiae*, and ITN would be expected to be even more effective with this species since it feeds at times when humans are most likely to be in bed. Admittedly, ITN and IRS would tend to produce some decrease in the abundance of *An. arabiensis*, but this is likely be relatively slight and offset in some degree by a reduction in interspecific competition. For example, Kirby & Lindsay (2009) demonstrated that immature stages of *An. arabiensis* survive better in the absence of *An. gambiae*. It is to be expected, therefore, that the reduction in numbers of *An. gambiae*, consequent on the increasing use of ITN, might even have resulted in an increase in populations of *An. arabiensis*.

### Analysis of the time course of malaria incidence

The spirit of the analysis developed below is that, in the first few years after the introduction of ITN, there was little if any insecticide resistance and most of the transmission of malaria was due to mosquitoes like An. *gambiae*, which feed primarily off humans, indoors and at night. Thereafter, transmission was due to mosquitoes like *An. arabiensis*, which have contrasting behaviour, and was aggravated by increased resistance to insecticides.

In keeping with these ideas, the WHO incidence data for Kenya and Tanzania (WHO, 2021a) were found to be well fitted by an initial exponential decline in infections, followed by a logistic increase (Fig. 5A, C):

**Figure 5.**
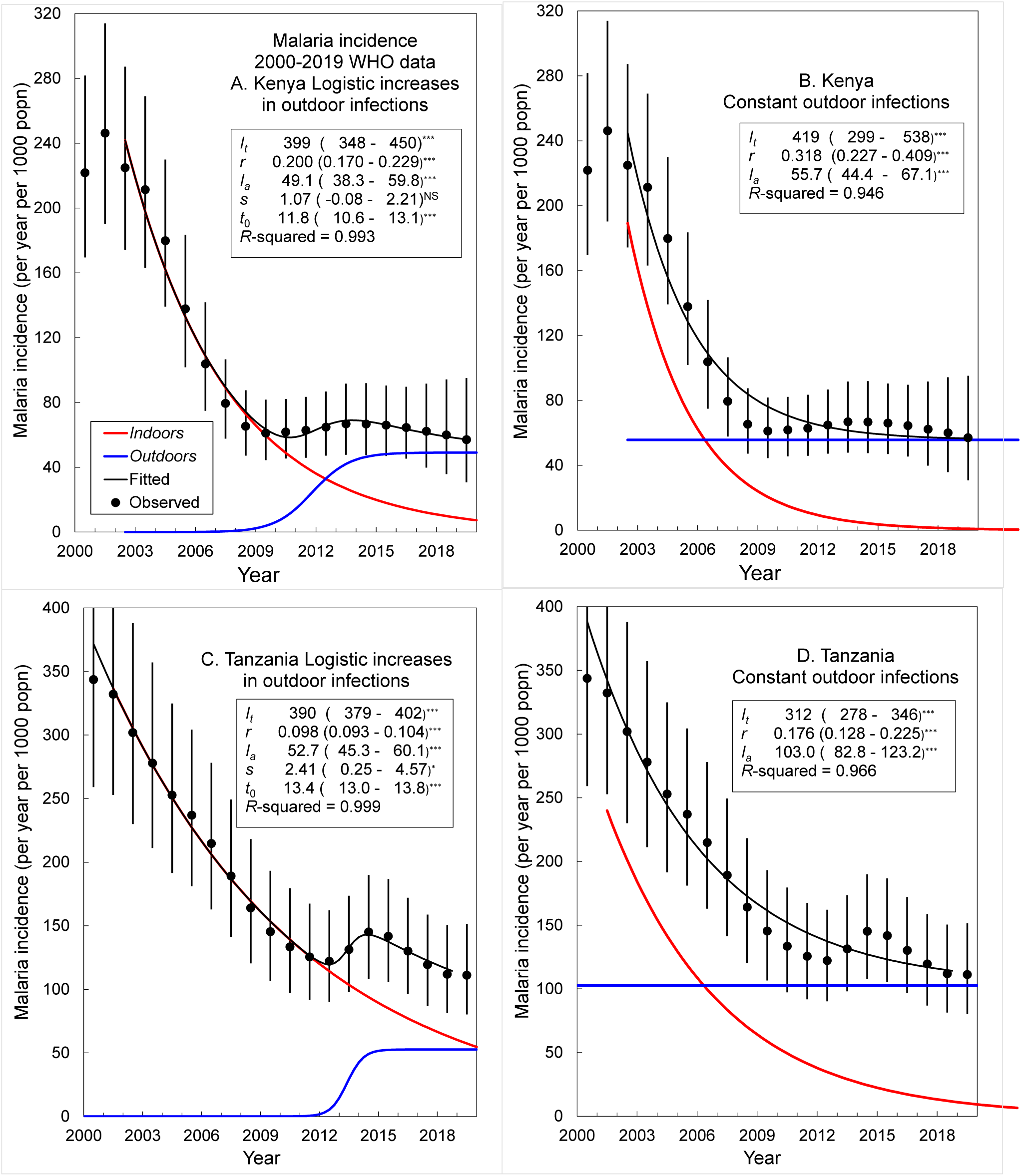
Modelling changes in malaria incidence in E Africa as the sum of infections incurred indoors, which declining exponentially with time, and incurred outdoors which either increase logistically with time, or remain at a constant level. The models are defined by Equation (1). Parameter values and 95% confidence intervals, estimated by non-linear least squares regression, are shown in the body of each graph. Key: ^***^ *P* < 0.001, ^*^ *P* < 0.05, ^NS^ *P* > 0.05.

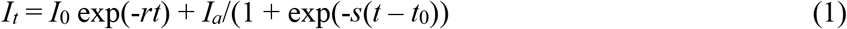

The first term describes the exponential decline, at rate *r*, from an initial incidence *I*_0_. The second term describes the logistic growth at rate *s* to an asymptotic level *I*_*a*_: *t*_0_is the value of *t* when the function reaches 50% of its maximum level.

Notice that the exponential decline in transmission was slower in Tanzania than in Kenya, presumably reflecting a relatively slow reduction in the abundance of endophagic mosquitoes and perhaps a more gradual development of insecticide resistance. In any event, the slow exponential decline in Tanzania fits with the fact that the logistic rise in transmission there occurred a few years later than in Kenya.

Other interpretations could be placed on the results. For example, the behavioural changes could be due to the selection of behaviourally distinct strains of *An. gambiae*, rather than adjustments in species balances We note, however, that an altered species balance fits better with the observed massive decline in the number of *An. gambiae*, and the large increase in the relative proportions of *An. arabiensis*. It is also possible to analyse the data assuming a constant component in the incidence of malaria (Fig. 5B, D). For this we take *s* = 0 in the second term of Equation (1), which then reduces simply to *I*_*a*_. Notice that this simplification reduces a little the goodness of fit.

### Outputs of the M model

We first produced simulations to compare the general ways that insecticide resistance and mosquito species govern the extents to which various levels of ITN ownership affect the abundance of mosquitoes and the incidence of malaria. We then made the simulations more specific by trying various ways of simulating the time-course of control by ITN in W Kenya, and then predicting the efficacy of supplementing ITN with either IRS or other control options there.

#### General simulations

Outputs for *An. gambiae* or *An. arabiensis* showed that nets that killed arriving mosquitoes could be far more effective than non-killing nets for malaria control, especially when many humans owned nets (Fig. 6). The results confirm that putting insecticide on the nets was a game-changer in malaria control. However, in the present context, the important corollary is that quite small reductions in kill rate, associated perhaps with insecticide resistance, can cause relatively large increases in the incidence of malaria. Such effects are as predicted by Ross (1911). They occur because the greater the abundance of mosquitoes the greater the number of infective bites the humans receive. That means that more humans are infected, which ensures that a higher proportion of mosquitoes get infected, thus enhancing even more the numbers of infective bites received by humans. Common-sense exceptions to this occur when anthropophilic mosquitoes are so numerous that humans get many infective bites per day, thus ensuring that even substantial changes in abundance cannot affect the incidence. However, it seems that mosquitoes were not so super-abundant in W Kenya in 2000-2004 since the reduction in their numbers, presumed to occur due to moderate levels of ITN ownership in the few years thereafter, was associated with a clear reduction in malaria incidence.

**Figure 6.**
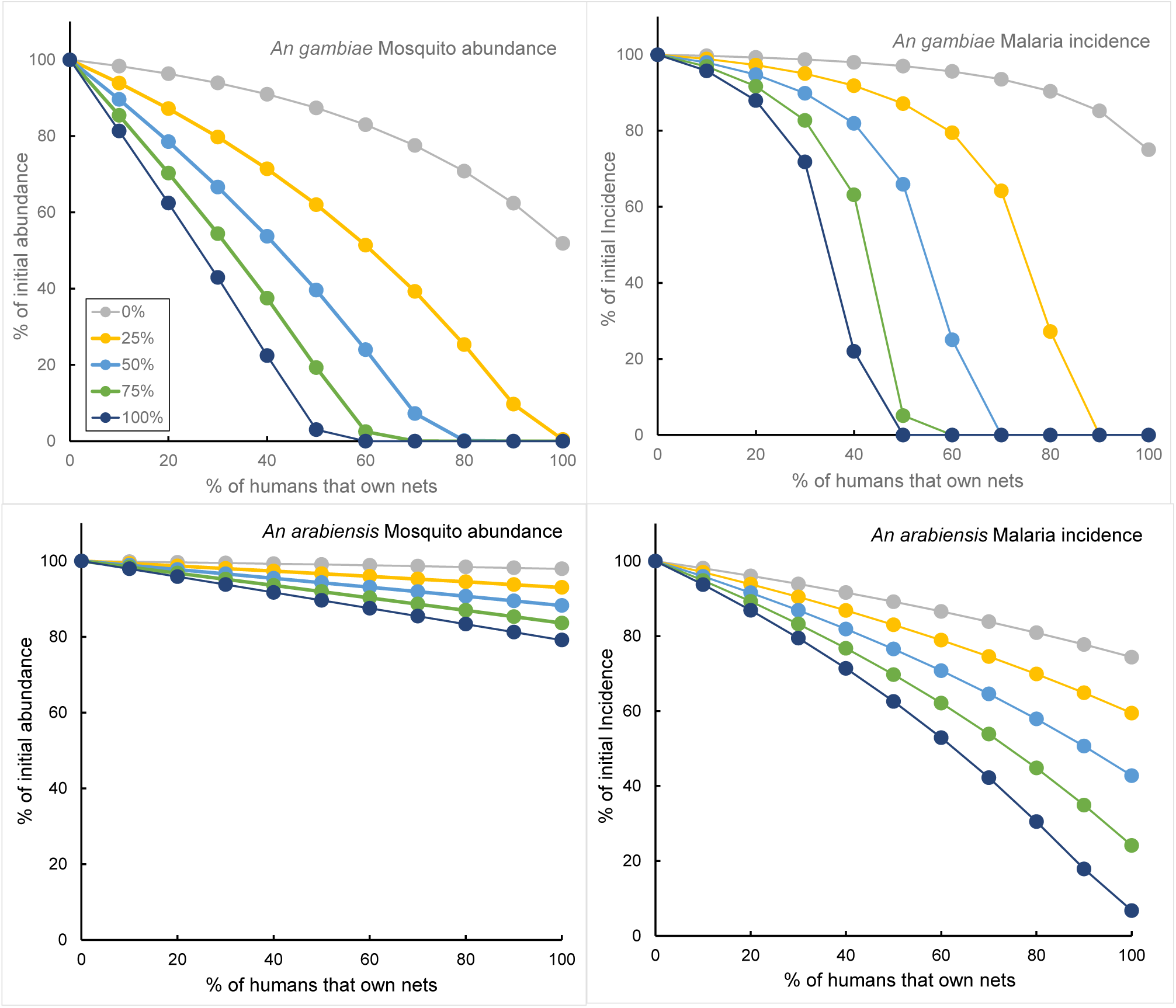
Average daily abundance of adult female mosquitoes and average daily incidence of mature infections of malaria (Y axis) in the fifth year of control campaigns employing ITNs against either *An. gambiae* (A) or *An. arabiensis* (B). The campaigns involved different percentages of net ownership (X axis) and various kill percentages (shown in legend). Averages are expressed as percentages of the average in the year prior to the start of control. In that year the average daily abundance of mosquitoes was 24,548/km^2^ for each species. The average incidence of mature infections was 4.67/ km^2^/day with *An. gambiae* and 2.46/ km^2^/day with *An. arabiensis*.

Comparison between the results for *An. gambiae* and *An. arabiensis* indicated that an ITN campaign of any given specification is markedly more effective for the former species (Fig. 6). For example, with net ownership of 50% and with insecticide deposits that killed 75% of the mosquitoes arriving at the nets, the results for *An. gambiae* show that the abundance of mosquitoes dropped to about 20% and malaria was virtually eliminated. For *An. arabiensis*, the mosquito abundance dropped to about 90% and the incidence of malaria declined to about 70%, *i*.*e*., the mosquitoes and malaria were far short of elimination. The salient point is that, where the two mosquito species are sympatric, the use of ITNs will be sufficient to reduce drastically the numbers of *An. gambiae* relative to *An. arabiensis –* even with moderate levels of bed-net ownership. We note, however, that – if it were possible to increase net ownership to 100%, with high levels of proper deployment of these nets – then the modelling suggests that malaria incidence due to *An. arabiensis* would also approach elimination (Fig. 6D).

#### Specific simulations

As already explained, it is currently impossible to model a situation involving a dynamic equilibrium between the many populations of sympatric mosquitoes in W Kenya. Hence, we cannot offer outputs suggesting how the *relative* abundance of *An. gambiae* changed. Against this, we could assess, as below, the credibility of the null hypothesis that the relative abundance of *An. gambiae* did *not* change in the whole period from 2000 to 2021. We could also examine how the development of insecticide resistance could have affected the outcome.

We recognised that our outputs should meet three criteria if they were to simulate well the observed pattern of malaria shown in Fig. 5A. First, there should be a sharp decline in the incidence of malaria between 2005 and 2009. Second, the decline must stop fairly sharply in 2010-2011, and third, the incidence from 2011 onwards must remain at roughly 25-30% of the level prior to control. In trying to satisfy these criteria we produced a sequence of simulations. For the initial group of simulations, the percentage kill at the nets was kept constant throughout the whole modelled period, as would be expected if no insecticide resistance developed. The results of this group (Fig. 7A) show that a constant kill rate of 70% produced a suitable decline in the incidence of malaria from 2005 to 2009 and was associated with an appropriate check in decline from 2010 to 2011. However, immediately after this, and in marked contrast with reality, the incidence of malaria dropped sharply to zero. This was true even when the kill rate was reduced to 60% or 50%. The only way of ensuring that the incidence did not decline to zero was to reduce the kill rate even more, but that resulted in an unrealistically slow decrease in incidence between 2005 and 2009. The next group of simulations attempted to avoid the occurrence of zero incidence by allowing for the known development of insecticide resistance after about 2004 (Ranson & Lissenden, 2016), presumably caused by the increase in ITN ownership and the consequent intensification of exposure to insecticide. It was taken that the kill rate was constant at 70% up to the end of 2004 and then declined linearly (loc. cit.) to the end of 2020. The results (Fig. 7B) show that if the kill rate declined to either 10% or 20% there was, as required, a stalling from 2010 to 2011, and persistent incidence thereafter. However, those two simulations were hardly satisfactory because the initial decline in incidence was not particularly marked, and especially because the incidence fluctuated greatly in the period of stalled control. Such fluctuations are hardly surprising given the great oscillations taken to occur in net ownership (Fig. 2A).

**Figure 7.**
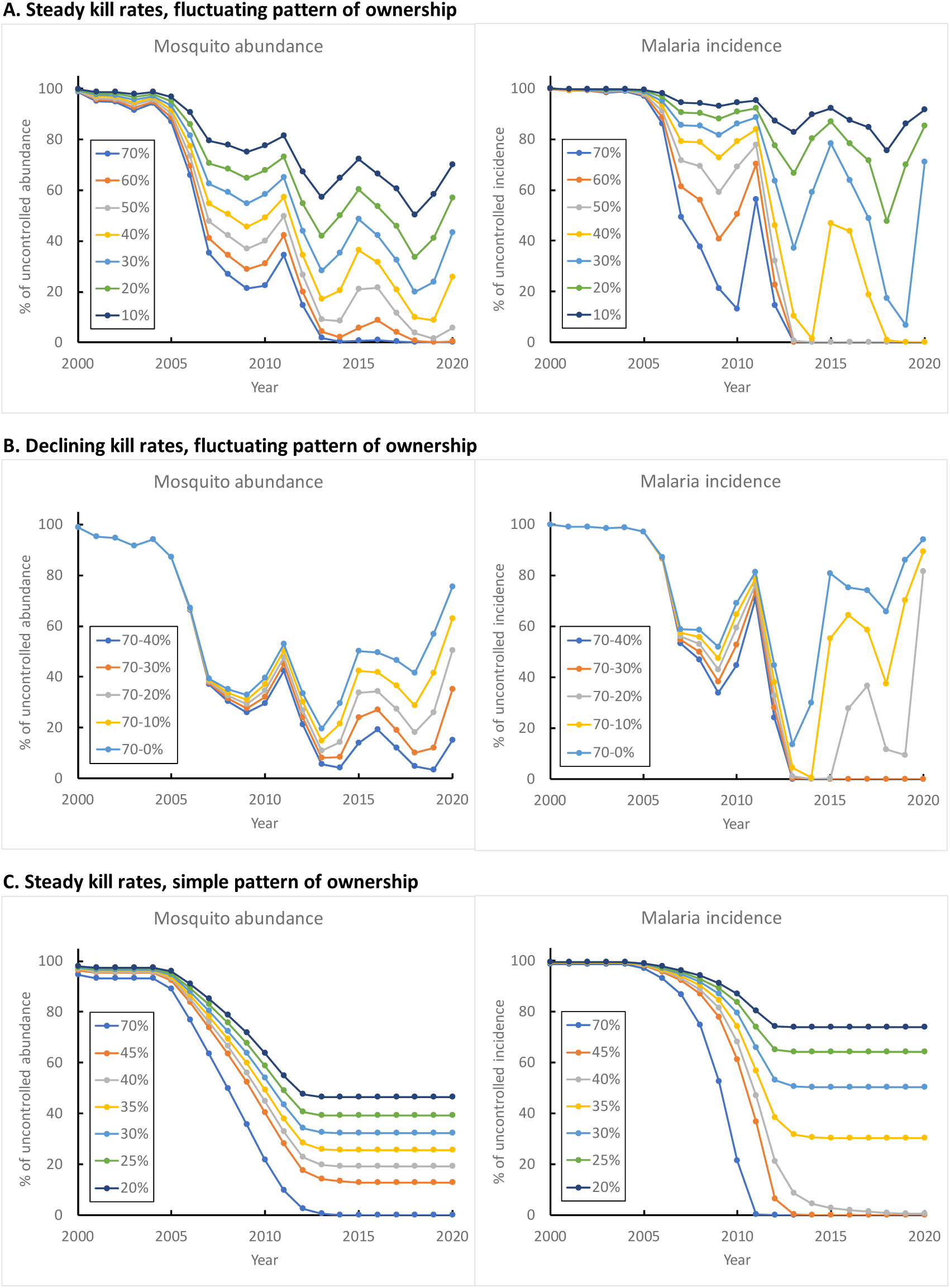
Simulated abundance of adult female mosquitoes and incidence of malaria (Y axis), in each year from 2000 to 2021 (X axis), expressed as a percentage of the uncontrolled abundance and incidence for 1999. In A the ownership of bed-nets fluctuated as shown in Fig. 2A; the kill rate at the nets (Figures in legend) was kept steady throughout all years of each simulation but was varied from 20% to 70% within different simulations. In B the ownership fluctuated as in A: the kill rate was always 70% from 2000 to 2004, but declined linearly thereafter, to become 0% to 40% in different simulations. In C the ownership followed the relatively simple pattern described in the text; the kill rate was steady within any simulation but varied between simulations. All simulations of A, B and C were performed on the hypothesis that the mosquitoes present were the standard strain *An. gambiae, i*.*e*., strongly anthropophilic and endophagic.

For the third group of simulations we tried to keep the incidence of malaria steady in the stalled period. For this we recognised the warning of Bertozzi-Villa et al. (2021) that their published time-course of net-coverage in Kenya was approximate, so that the fluctuations in coverage might be excessive. Hence, we assumed that the ownership of nets varied in a relatively simple way, without oscillations. Ownership was assumed steady at 5% from 2000-2004, rising linearly to 70% during 2005 to 2012, and then remaining steady at 70% up to the end of 2021. In further avoidance of fluctuating incidence, the percent kill at the nets was also steady throughout each simulation but was varied from 20% to 70% within different simulations. The effective use of owned nets was kept steady throughout at the 75% adopted in previous simulations with *An. gambiae*. The results (Fig. 7C) showed that the incidence of malaria could be suitably level in the stalled period, provided the kill rate was not too high. A kill rate of 35% ensured that the incidence in the stalled period was at the realistic level of 30%. However, with that degree of percentage kill the simulation was unsatisfactory in that initial decline in incidence was unrealistically slow and the stalled control emerge too late, by 3-4 years.

The above groups of simulations, based on the hypothesis that *An. gambiae* predominated throughout, indicate the impossibility of getting a simple, confident simulation of the required pattern of malaria control. Thus, with no resistance (Fig. 7A) there would have been no stalling in malaria control. If resistance did develop (Fig. 7B) there could easily have been stalling, but substantial fluctuations in incidence would have been likely to occur. To avoid fluctuations in incidence, it would be easiest to envisage no oscillation in net ownership (Fig. 7C) but then stalling would occur too slowly. Admittedly, if one were to abandon the concept of relatively gradual changes in the parameters governing control, it would be possible to engineer simulations that were close to reality. For example, in Fig. 7C, one could imagine that the incidence curve for a 70% kill rate applied initially, and then was suddenly replaced by a steady kill rate of 35%. However, it seems that any way of simulating the W Kenya experience would point to something(s) being awry in the efficacy of ITN in the latter half of the modelling period. Hence, if we still maintain that the majority of mosquitoes were anthropophilic and endophagic, we would have to suppose that the problem was with the way in which the ITN was applied. For example, the nets were not being distributed fully or used properly, or they were seriously degraded, or their insecticide was poorly toxic. The implication of such eventualities would be that IRS employed on top of ITN from 2017 onwards would have had plenty of scope for enhancing the overall degree of control – especially since it is well established that the organophosphate insecticide used for the IRS covered 92% of the dwellings (PMI, 2021) and was highly toxic to mosquitoes (Animut & Horstmann, 2022). In the next section we present results showing the large difference between the expected levels of control due to IRS, and the levels of control observed in the field.

#### Addition of IRS

To consider how effective IRS should have been when imposed in addition to ITN, it was allowed that net ownership was constant at 70% and the kill rate was a steady 35%. This ensured that in the absence of IRS the incidence of malaria would be stalled at a fixed 30% of the pre-control level (Fig. 7C). Taking the IRS coverage at a conservative 80%, our modelling suggested that the IRS, when added to the use of ITN, would have resulted in the virtual elimination of malaria in a year, and even the very low rate of 5% coverage would have caused the incidence of malaria to drop to only about 10% of the initial 1999 level in four years (Fig. 8).

**Figure 8.**
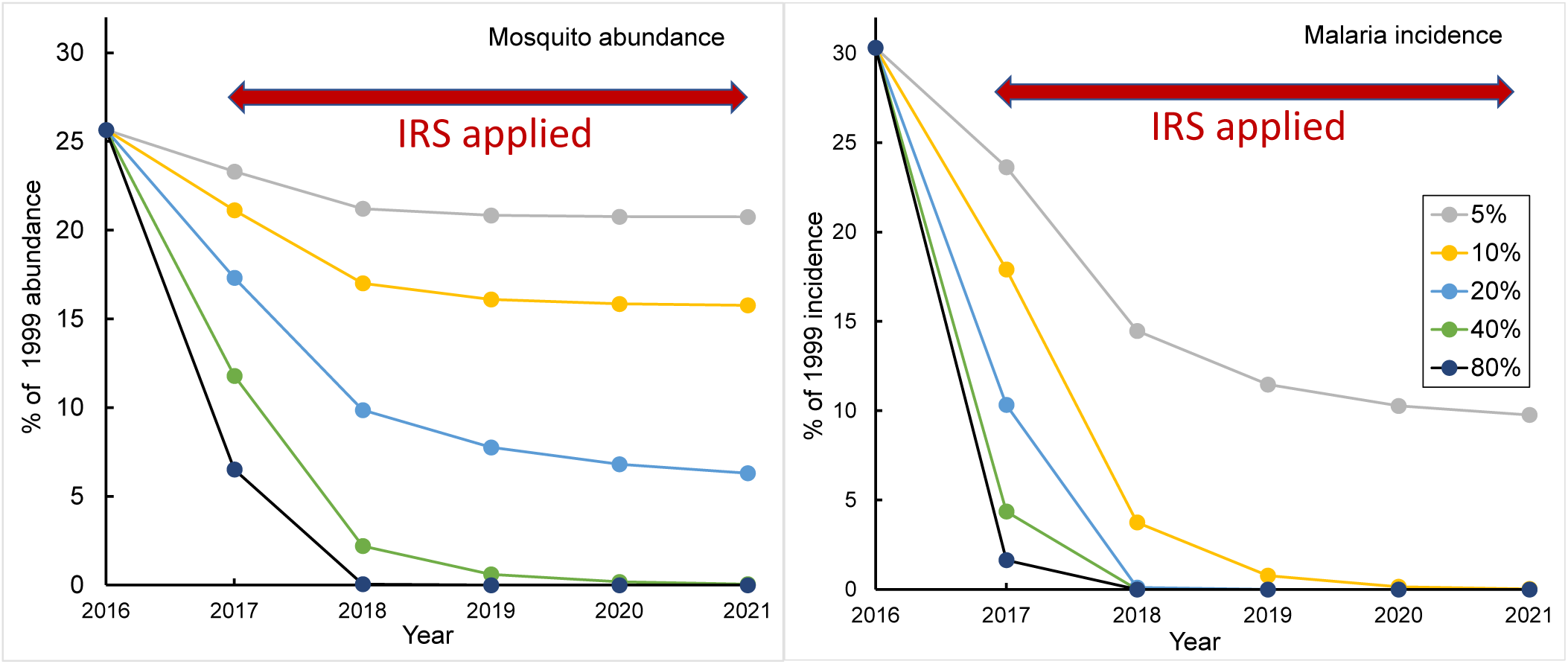
Simulated effects of IRS on the abundance of adult female *An. gambiae* and the incidence of malaria (Y axis), in each year from 2016 to 2021 (X axis), expressed as a percentage of the uncontrolled abundance and incidence for 1999. The IRS was applied from 2017 to 2021, at various percentage coverages of dwellings (numbers in legend). It was taken that 75% of the mosquitoes fed indoors and that the IRS insecticide killed 75% of the mosquitoes so feeding. The use of insecticide treated nets was applied throughout, with 70% ownership of nets and a kill rate of 35% at the nets

Summarising the indications of the simulations, it seems impossible to maintain the hypothesis that *An. gambiae* predominated throughout. The implication is that an exophagic vector such as *An. arabiensis* moved centre-stage. We certainly do not say that insecticide resistance has been unimportant, nor that bed-nets have not played a very significant part in reducing incidence and saving lives. However, the more that exophagy has developed the less the potential efficacy of indoor campaigns, so that the less has been the impact of changing the numbers of bed-nets and the toxicity of the insecticide they employ.

#### Control of zoophilic and exophagic mosquitoes

Allowing that mosquitoes such as *An. arabiensis* are not being fully controlled by ITN and/or IRS in E Africa, it is necessary to consider the efficacy of additional control methods that could be more suited to zoophilic and exophagic species. In particular, it is required to consider controlling the mosquitoes in their aquatic stages or while they are visiting non-humans. Simulations employing these two types of control with *An. arabiensis* indicate that, at any assumed degree of ITN ownership, the killing of mosquitoes visiting non-humans tends to be much more effective than the reduction in adult emergence (Fig. 9). This is because the reduction in emergence is applied once-off per life cycle of many days, whereas the opportunity to kill the feeding flies occurs once every few days of the gonotrophic cycle. It is especially noteworthy that a kill rate of just 10% per feed on non-humans led to the elimination of malaria, even when there was no IRS, and the concurrent ownership of bed-nets was no more than the 40% shown by field experience to be well within achievable limits. It follows from Fig. 9 that, if the ownership of bed-nets were increased close to 100%, then malaria could be eliminated by kill rates of markedly less than 10% at non-humans.

**Figure 9.**
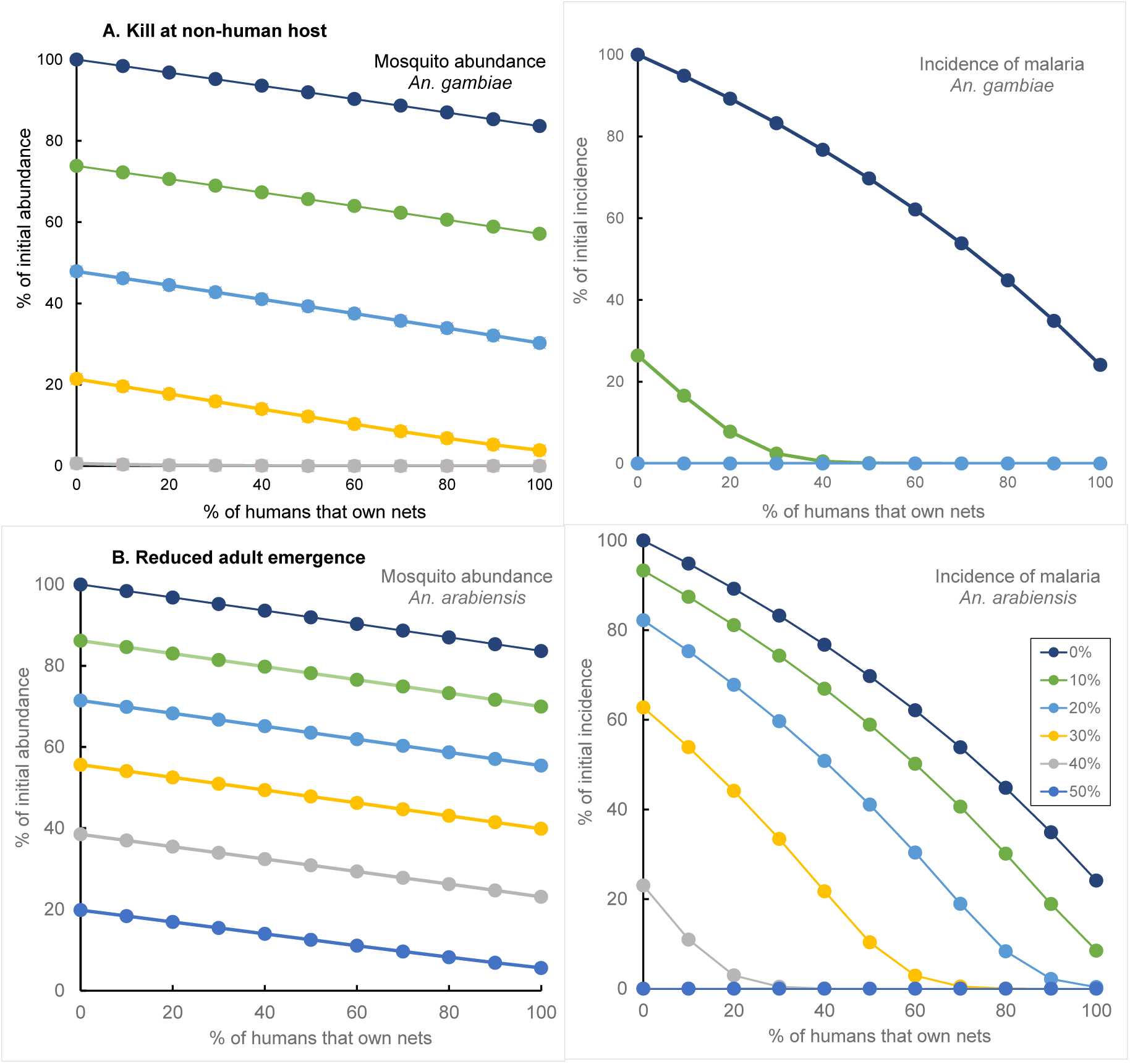
Simulated abundance of adult female *An. arabiensis* and the incidence of malaria (Y axis), resulting from five years of control by killing adults that arrived at non-humans (A) or by reducing the daily rate of adult emergence (B). Abundance and incidence are expressed as percentages of their initial, uncontrolled values. The use of insecticide treated nets was applied with various percentages of bed-net ownership (X axis) but always involved a 75% use of nets and a kill rate of 75% for mosquitoes arriving at the nets. Figures in the legends refer to the percent kills or percent reductions in emergence.

## Discussion

### Literature search

While the literature search was primarily intended to indicate the time-course of control measures and their effects in W Kenya, in preparation for our modelling of that situation, it emerged that field experiences there matched those elsewhere. For example, the long-term effects of indoor application of insecticides seem to have resulted in shifts towards daytime feeding outdoors by: (i) *An. gambiae* in Equatorial Guinea (Reddy et al., 2011); (ii) *An. gambiae* and *An. funestus* in Tanzania (Russell et al., 2011), and (iii) *An. funestus* in Senegal (Sougoufara et al., 2014). Moreover, in analysing data from experimental huts in Tanzania, it was noted that: “ITNs treated with pyrethroids were more effective at killing *An. gambiae* and *An. funestus* than *An. arabiensis*. This could be a major contributing factor to the species shifts observed in East Africa … (and) *An. arabiensis* is likely to remain responsible for residual malaria transmission … over larger areas” (Kitau et al., 2012). Many other publications highlight these matters and frequently state the consequent need for methods of vector control other than ITN or IRS (Kitau et al., 2012; Durnez & Coosemans, 2013; Killeen, 2014; Sougoufara et al., 2014; Njoroge et al., 2017; Abong’o et al. 2018, 2020; Loha et al., 2019; Degefa et al., 2021; Lindsay et al., 2021).

#### Mathematical analyses and M modelling

The analyses and modelling were intended to offer quantitative support to the above considerations We do not attach great importance to the good fit of our mathematical model to the WHO data for the incidence of malaria (Fig. 5) – particularly since those data are themselves the product of a model (WHO, 2021a; World Malaria Report, 2021), which used in turn the net access and usage estimates of Bertozzi-Villa et al (2021), which were themselves the output of a modelling process. The important features of the data, which appear to be generally accepted (Bhatt et al., 2015), are simply the rapid decline in malaria incidence in the first decade of the century – and the lack of material change thereafter, in the face of continued high and often increasing use of ITN and in some instances the use of intensive IRS.

Moreover, we accept that the results of the M modelling should be regarded as illustrating quantitative principles, rather than offering precise predictions. For example, consider the outputs indicating that malaria transmitted by *An. arabiensis* can be eliminated by killing 10% of the mosquitoes feeding on non-humans, coupled with a 40% ownership of bed-nets. This should be taken as suggesting no more than the likelihood that modest levels of effective insecticide treatment of non-humans, coupled with realistic degrees of bed-net ownership, could overcome much or most, even all, of the stalled control of malaria in W Kenya.

The potential benefit of reducing the emergence rate of adults is well illustrated by the fact that such control eliminated *Anopheles* populations in Cyprus (Shelley & Aziz, 1949) and Brazil (Killeen et al., 2002). The field evidence for the efficacy of insecticide treatment of non-humans might seem less encouraging – it being indicated that the application of deltamethrin to cattle had no convincing impact on mosquito abundance in a small pilot study in W Kenya, largely because the insecticide was not suitably persistent (Njoroge et al., 2017). However, the first small trials of insecticide-treated cattle to control tsetse flies were also regarded as disappointing (Whiteside, 1949) before further research led to its improvement and widespread incorporation into successful strategies of control (Torr et al., 2005).

#### Future work

There is a need to confirm with other models the main indications of the present work, and to evaluate in fuller detail and confidence the many sorts of inputs required for the modelling of various control options. Perhaps the most exciting option is the possibility of a field trial where increased ITN coverage is combined with treatment of mosquitoes visiting non-humans outdoors. We suggest field testing of combinations of the following interventions: (i) increasing ITN coverage until it is as close as possible to 100%; (ii) introducing objective monitoring systems to maximise effective use of these nets (Krezanoski et al., 2017), and (iii) treating livestock with chemicals such as ivermectin and fipronil (Makhanthisa et al. (2021).

It is encouraging that there is already a rising interest in refining the efficacy of insecticide-treated livestock (Ruiz-Castillo et al., 2021), involving for example the use of endectocides, which might be more suitable than the pyrethroids used in the earlier work in W Kenya (Njoroge et al., 2017). Moreover, existing modelling has already predicted that the use of endectocides on cattle can be a highly effective complement to the use of ITN (Yakob et al., 2016). Laboratory trials suggest that a combination of injected ivermectin and topically applied fipronil could increase mortality in *An. arabiensis* by 2.4 – 3.5-fold and reduce fecundity by 60-90% (Makhanthisa et al., 2021).

Experience in the development of bait technology for tsetse (Vale & Torr, 2004) offers some suggestions for important principles to adopt more fully with mosquitoes. For example, with tsetse it has been shown that artificial baits can be highly effective where there are insufficient cattle. Moreover, artificial baits are of interest because the pyrethroids deposited on the fabrics of such baits can remain effective for at least a year (Mangwiro et al., 1999), as against only a few weeks on cattle (Njoroge et al., 2017). Irrespective of what baits are used, however, it is required to know the absolute density of mosquito populations, and what catch levels mean about that density, since this allows better predictions for the impact of bait control. It is also necessary to know how many of the mosquitoes that visit baits are actually caught or killed by them, because this exposes the scope for improving efficiency. Understanding the distance at which wild-bred and unrestricted mosquitoes can sense and respond effectively to sources of attractive stimuli is helpful in assessing bait performance. Knowing the distances that mosquitoes move during the totality of their activities is crucial in planning the scale of bait deployment and in countering reinvasion of treated areas.

We acknowledge that the above sorts of research take a long time, so we do not suggest that field application of existing outdoor techniques should be delayed until the research is complete. Moreover, we caution that the use of any outdoor technique should not be accompanied by a relaxation in commitment to ITN and IRS. Such relaxation may be expected to produce a resurgence in numbers of endophagic mosquitoes.

#### Conclusions

Malaria control has been stalled now for more than a decade and continued exclusive use of current levels of IRS and ITN will not result in any significant improvement. If, however, ITN ownership can be increased closer to 100% – with high levels of effective use of those nets, even quite modest levels of additional control outdoors should result in substantial reductions in incidence, even if they do not eliminate the disease. Treatment of adult mosquitoes visiting livestock and/or artificial baits seems the most promising of the outdoor options.

## Data Availability

All data produced in the present study are available upon reasonable request to the authors

https://drive.google.com/drive/folders/1Vwqz0rCtpMo775ePf-Qv10Q8FaPA5rSw?usp=sharing

## Abbreviations

ATSB: attractive toxic sugar baits
ITL: Insecticide treated livestock
ITN: insecticide-treated bed-nets
LLIN: Long-lasting insecticidal nets
IRS: Indoor residual spraying
KHIS: Kenya Health Information System
LSM: larval source management
PMI: United States President’s Malaria Initiative
WHO: World Health Organization.

## Acknowledgements

We are extremely grateful to Daniel Mwai for malaria incidence data extracted from KHIS records, to Amelia Bertozzi-Villa for providing us with full access to data on estimates of the distribution and usage of insecticide treated nets, and to Abdisalan Noor for details of the procedures used to produce WHO’s malaria incidence estimates. We thank Carlo Costantini, Gay Gibson, Jo Lines, Stephen Lindsay and Stephen Torr for extensive discussions of, and suggestions for, this paper.

## Authors’ contributions

GAV developed the Excel model; JWH developed the mathematical analyses. Both authors contributed equally to all other aspects of the paper.

## Availability of data and materials

The Excel model used in this study can be accessed at ***********. The authors generated no new data of their own. Data that support the findings of this study, accessed from the third parties named in the Acknowledgement, are available from those third parties. All materials generated in this publication are available from the authors on request.

## Declarations

### Ethics approval

Not applicable; the study involves theoretical modelling, and analysis of published data.

## Consent for publication

Both authors gave consent for this publication.

